# Nucleus Accumbens-Anterior Limb of Internal Capsule Deep Brain Stimulation for Treatment-Resistant Schizophrenia: An Open-label Pilot Study

**DOI:** 10.64898/2026.10.01.26364314

**Authors:** Chang Lu, Xin Zhang, Zhaolin Zhai, Jingxin Xue, Yuqing Zhao, Qiong Xiang, Kaiming Zhuo, Yifeng Shen, Huiqin Han, Hongbing Lei, Peng Wang, Lina Shen, Liqin Lang, Lei Chen, Yifeng Xu, Zhen Wang, Dengtang Liu

## Abstract

We conducted an open-label pilot study of bilateral nucleus accumbens-anterior limb of the internal capsule (NAc-ALIC) deep brain stimulation (DBS) in three patients with treatment-resistant schizophrenia (TRS). Clinical symptoms, global functioning, cognition, stimulation parameters and intracranial local field potential (LFP) were assessed over 24 weeks. Two patients met the prespecified clinical response criterion, defined as a ≥40% reduction in Positive and Negative Syndrome Scale (PANSS) total score, by week 8 and showed PANSS reductions of 69.0% and 69.4%, respectively, at week 24. The third patient showed a 18.7% reduction at week 24. Responders showed improvements in global functioning and selected cognitive domains. Longitudinal LFP recordings revealed increased gamma-band relative power and reduced theta-band activity in responders, whereas these changes were less consistent in the non-responder. DBS was generally well tolerated, with transient stimulation-related adverse effects resolving after reprogramming. These findings provide preliminary clinical evidence supporting further investigation of NAc-ALIC DBS in TRS and suggest that individualized programming and intracranial physiological measures may contribute to the identification of treatment-response signatures.

**Registration:** NCT06112067 in ClinicalTrial.gov

## Introduction

Deep brain stimulation (DBS) is an established neuromodulatory intervention for several neurological disorders, particularly movement disorders such as Parkinson’s disease, dystonia, and essential tremor, and has increasingly been investigated for neuropsychiatric disorders^1^. DBS involves the stereotactic implantation of electrodes to deliver electrical stimulation to anatomically defined brain regions. Although DBS was initially conceptualized as producing a local inhibitory effect, accumulating experimental evidence indicates that its therapeutic effects cannot be explained by simple focal inhibition alone and may instead involve modulation of pathological activity across interconnected neural circuits^2,3^. Unlike irreversible ablative procedures, DBS permits adjustable and potentially reversible neuromodulation, allowing stimulation parameters and active contacts to be tailored to individual clinical response and tolerability.

Among psychiatric disorders, DBS has been most extensively investigated in obsessive-compulsive disorder (OCD) and treatment-resistant depression (TRD)^4^. For severe, treatment-refractory OCD, stimulation of the ventral capsule/ventral striatum, including the nucleus accumbens (NAc), has been investigated in some clinical studies^5,6^. For TRD, the subcallosal cingulate cortex and related fronto-limbic circuits have emerged as prominent targets^7,8^. By comparison, the therapeutic potential of DBS for treatment-resistant schizophrenia (TRS) remains largely unexplored. A limited number of clinical studies and case reports have investigated DBS at various targets. In a pilot randomized crossover trial by Corripio et al two of three patients receiving NAc stimulation and two of four receiving anterior cingulate stimulation met the prespecified response criterion, providing preliminary evidence that DBS may ameliorate persistent psychotic symptoms in a subset of patients^9^. Subsequent small studies have reported clinical improvements following habenula DBS in two patients^10^ and substantia nigra pars reticulata DBS in an individual with TRS^11^.

Schizophrenia has consistently been associated with abnormalities in emotional processing and motivational drive, suggesting a disruption in the integration of affective experience with goal-directed behavior^12^. The NAc represents a plausible target for circuit-based intervention in schizophrenia at anatomical, functional, and network levels^13,14^. Anatomically, the NAc lies at the convergence of prefrontal cortex, hippocampal, and amygdala inputs, and serves as a key node in the processing of motivation and salience^15^. Its shell and core subregions receive partially distinct limbic and motor-related inputs, respectively, positioning the NAc at a crucial interface between motivational, emotional, and action-related circuits^16^. At the neurochemical level, preclinical studies suggest that NAc DBS can alter ventral tegmental area (VTA)-related dopamine signaling^17^. Dysregulated dopamine signaling has been implicated in psychosis through its effects on prediction-error processing and the attribution of salience to otherwise irrelevant stimuli^18,19^. Experimental studies have further provided causal evidence linking increased striatal dopamine activity to hallucination-like perceptual experiences^20,21^. Contemporary models of schizophrenia similarly implicate dysregulated striatal dopamine signaling in psychosis, while emphasizing that this dysfunction is distributed across interconnected striatal circuits rather than a simple, anatomically restricted “mesolimbic dopamine excess”^22^.

The prevailing hypothesis for TRS pathogenesis is the excitation-inhibition (E-I) imbalance, characterized by dysregulated glutamate and gamma-aminobutyric acid (GABA) signaling originating from the striatum, which subsequently leads to downstream dopamine hyperactivity^23,24^. The NAc is predominantly composed of GABAergic medium spiny neurons^25^, and high-frequency stimulation of the NAc has been reported to modulate local neural activity and GABAergic signaling^26^. Importantly, any local neurochemical effects of NAc DBS are likely to propagate through broader dysfunctional networks implicated in persistent psychotic symptoms^27^. The anterior limb of the internal capsule (ALIC) provides a complementary anatomical target because it contains fronto-striatal and fronto-limbic projection fibers connecting prefrontal and limbic regions with subcortical structures. Alterations in the microstructural integrity of the ALIC have been reported in schizophrenia, further implicating this pathway in the disorder^28,29^. From a therapeutic perspective, stimulation encompassing the NAc and adjacent ALIC may therefore engage both a ventral striatal node and the fronto-subcortical fiber pathways. This NAc-ALIC configuration is also consistent with the broader circuit-based framework in which clinical benefit may depend on modulation of distributed networks rather than isolated nuclei.

Here, we conducted a prospective, open-label pilot clinical study of bilateral NAc-ALIC DBS in three patients with severe TRS. We longitudinally assessed psychopathology, global functioning, neurocognition, stimulation-related adverse effects, and individualized DBS programming trajectories over 24 weeks. In addition, LFPs were recorded during stimulation-off periods at baseline and follow-up visits to explore electrophysiological changes. By integrating clinical, programming and intracranial electrophysiological data, this study aimed to provide preliminary evidence for the clinical feasibility and tolerability of NAc–ALIC DBS and to generate hypotheses concerning neural signatures that may inform future biomarker-guided DBS strategies for TRS.

## Results

Between November 2023 and May 2025, 51 patients with schizophrenia were assessed for eligibility. Of these, 28 did not meet the inclusion criteria, three underwent diagnostic revision following clinical reassessment, and 14 declined participations, including three who chose to enroll in other clinical trials. Following written informed consent from both patients and their legal caregivers, three participants were ultimately enrolled. The study timeline is shown in Fig. 1a.

**Fig. 1.**
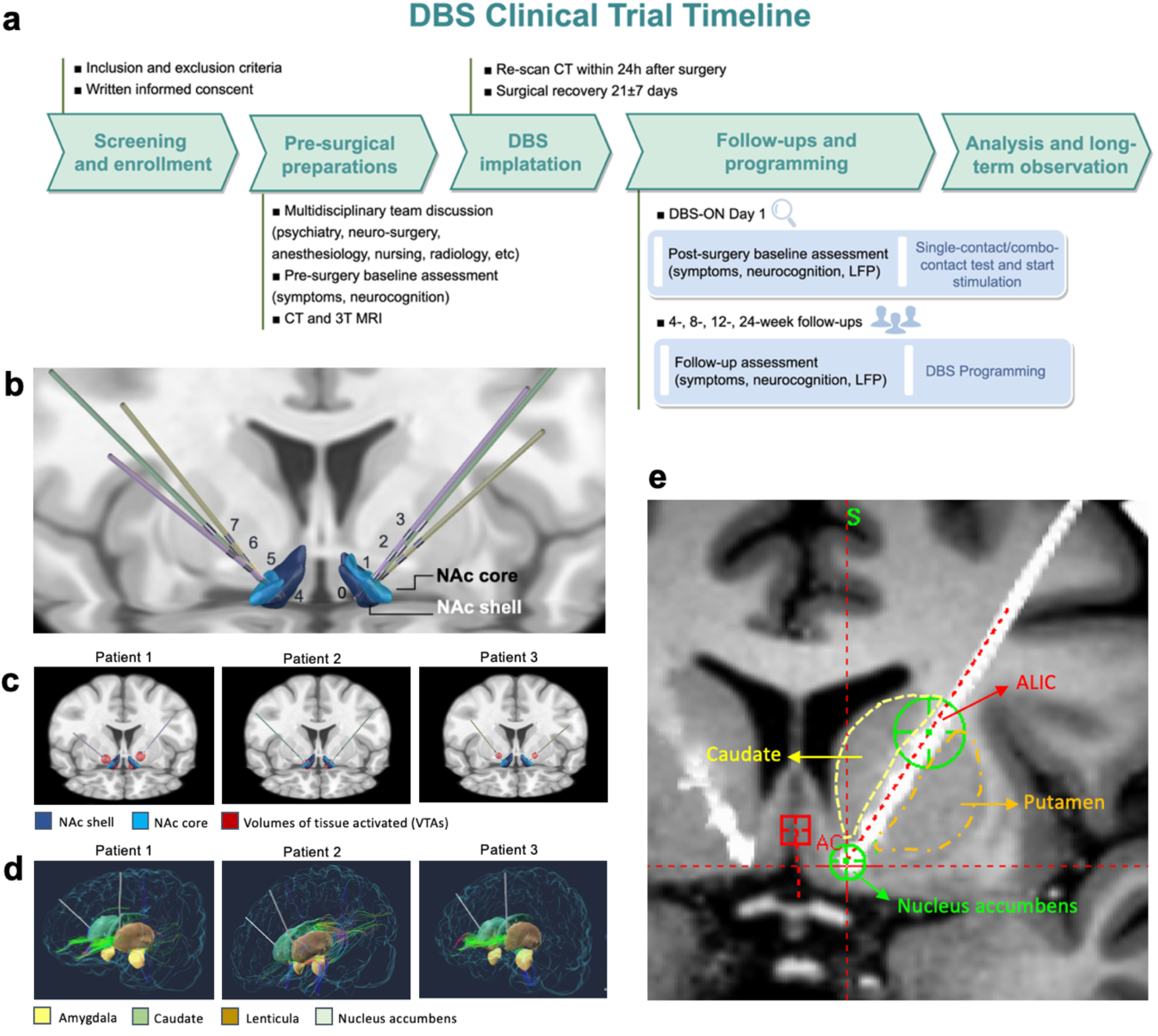
Study timeline, DBS lead localization, patient-specific VTAs, and connectivity. (a) DBS clinical trial timeline. (b) Three-dimensional reconstruction of bilateral DBS lead locations for the three enrolled patients in MNI space. Leads from Patient 1, Patient 2, and Patient 3 are shown in purple, green, and yellow, respectively. The NAc core and shell are shown in light and dark blue, respectively. (c) Patient-specific volumes of tissue activated (VTAs) reconstructed from the stimulation parameters selected during initial postoperative programming. The illustrated settings produced favorable acute symptom responses while maintaining acceptable tolerability during initial DBS programming. Stimulation parameters were as follows: Patient 1, contacts C0/C2/C4/C6, 4.5 V, 210 μs, 145 Hz; Patient 2, contacts C0/C4, 4.0 V, 210 μs, 130 Hz; and Patient 3, contacts C2/C6, 2.5 V, 210 μs, 130 Hz. (d) Patient-specific diffusion MRI-based tractography seeded from the VTAs, illustrating their structural projections to major brain regions implicated in the relevant neural circuits. (e) Preoperative T1-weighted MRI co-registered with postoperative CT in Patient 1. The lead trajectory and electrode configuration enabled simultaneous coverage of the anatomically adjacent NAc and ALIC.

### Demographic and preoperative characteristics

The three patients, aged between 22 and 29 years, exhibited illness durations ranging 7 to 14 years. Each had a history of multiple inadequate responses to antipsychotic medications and had previously undergone treatment with modified electroconvulsive therapy (MECT). At baseline, the severity of symptoms was considerable, as indicated by PANSS total scores ranging from 92 to 121, Clinical Global Impression–Severity (CGI-S) scores of 5-6, and markedly impaired functioning, as reflected by Global Assessment of Functioning (GAF) scores between 25 and 35.

At enrollment, all patients were receiving clozapine-based regimens, with one patient additionally receiving long-acting injectable paliperidone. Maximum clozapine doses were 300 mg/day, resulting in plasma concentrations of 383.0, 227.7, and 556.8 ng/mL for Patient 1, 2, and 3, respectively. All participants completed the 24-week follow-up without attrition. Reconstructed DBS electrode positions, volumes of tissue activated (VTAs), neuron projections and adjacent brain structures are shown in Fig. 1b-e, respectively. Prior to DBS implantation, antipsychotic treatment had remained stable for at least 2 months. No modifications to the type, formulation, or dosage of antipsychotics were made during the DBS intervention or the prespecified follow-up period.

### Clinical symptoms

At Week 12 after DBS initiation, the PANSS total score had decreased by 66.2% in Patient 1 (101 to 54), 15.4% in Patient 2 (121 to 107), and 48.4% in Patient 3 (92 to 62). By week 24, the corresponding reduction rates were 69.0%, 18.7%, and 69.4%, respectively.

Patient 1 and 3 met the prespecified clinical response criterion at Week 8 (Table 2, Fig. 2). Clinically significant improvements in both positive and negative symptoms were observed in both responders throughout study visits, including reductions in delusions, conceptual disorganization, emotional withdrawal, poor rapport, and apathy. Depressive symptoms assessed by the Calgary Depression Scale for Schizophrenia (CDSS), remained absent throughout follow-up in all three patients.

**Fig. 2.**
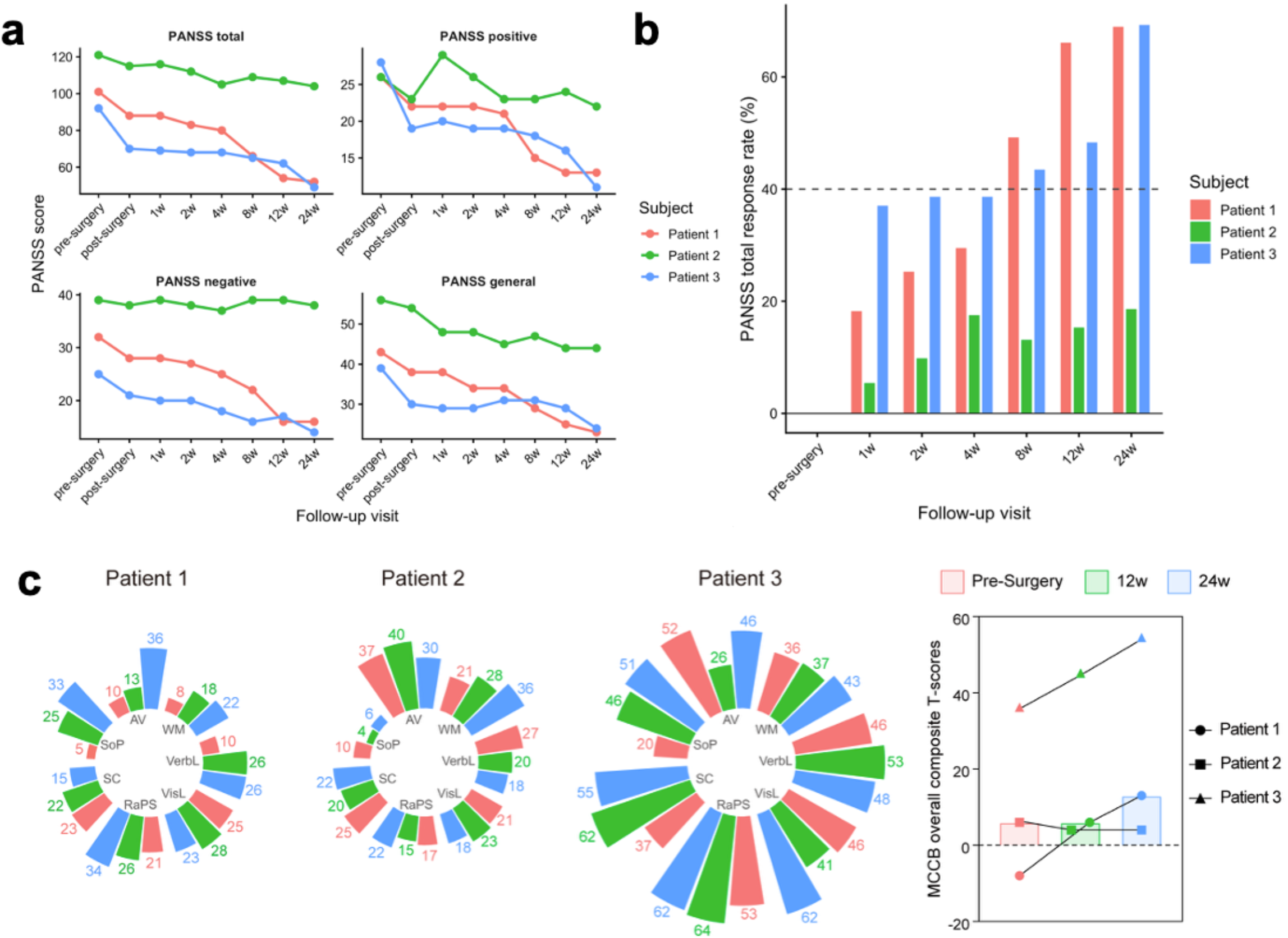
Clinical and cognitive outcomes during NAc-ALIC DBS treatment. (a) PANSS total scores and positive, negative, and general psychopathology subscale scores; (b) PANSS reduction rates; and (c) MATRICS Consensus Cognitive Battery (MCCB) composite T-scores. SoP, speed of processing; AV, attention/vigilance; WM, working memory; VerbL, verbal learning; VisL, visual learning; RaPS, reasoning and problem-solving; SC, social cognition.

**Table 1.** Patient demographic and pre-surgery baseline information.

| Patient | Patient 1 | Patient 2 | Patient 3 |
| --- | --- | --- | --- |
| Sex | Female | Female | Male |
| Schizophrenia illness duration (years) | 7 | 12 | 14 |
| TRS symptoms | Auditory hallucination, inappropriate laughter, incongruous affect, attention deficits | Persecutory delusion, referential delusion, auditory hallucination, verbal hostility, blunted affect | Persecutory delusion, auditory hallucination, verbal and behavioral impulsivity |
| Past MECT | Yes | Yes | Yes |
| PANSS total | 101 | 121 | 92 |
| CGI-S | 5 | 6 | 5 |
| GAF | 35 | 25 | 31 |
| Past failed antipsychotics | Risperidone, aripiprazole, amisulpride, quetiapine, blonanserin, clozapine | Amisulpride, paliperidone, aripiprazole, risperidone, olanzapine, perphenazine, clozapine | Risperidone, olanzapine, paliperidone, clozapine |
| Maximum clozapine dosage (mg/d) | 300 | 300 | 300 |
| Clozapine plasma concentration at Baseline (ng/ml) | 383.0 | 227.7 | 556.8 |
| Medication at enrollment | Clozapine 200mg/d | Clozapine 150mg/d | Clozapine 300mg/d + Paliperidone LAIs 150mg/4w |
| Comorbidities | None | Hepatic dysfunction (mild-elevated hepatic function test results) | None |

MECT, Modified Electroconvulsive Therapy; PANSS, Positive and Negative Syndrome Scale; CGI-S, Clinical Global Impressions–Severity scale; GAF, Global Assessment of Functioning; LAIs, long-acting injectables; TRS, treatment-resistant schizophrenia

**Table 2.**
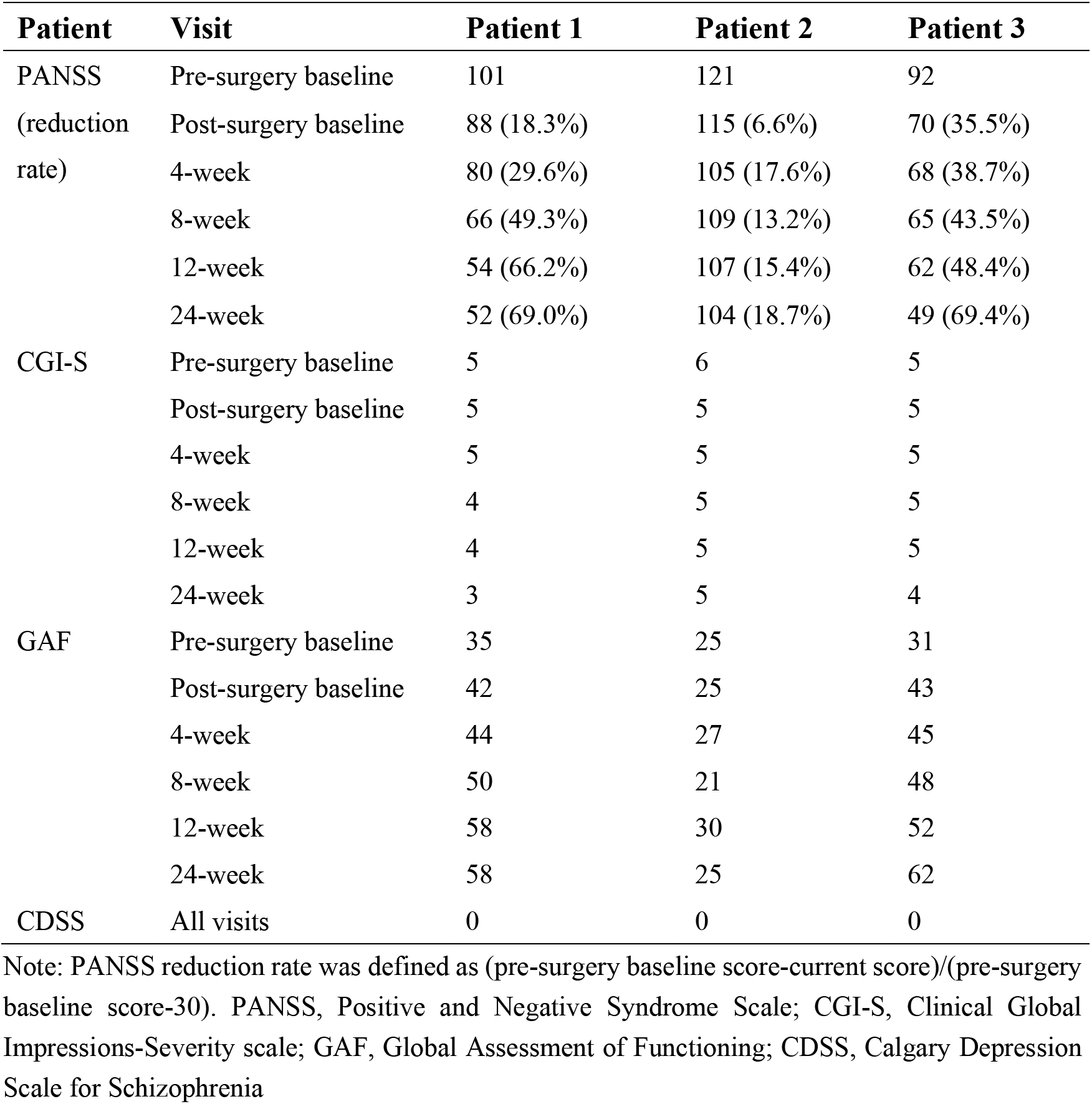
Longitudinal changes in PANSS, CGI-S, GAF and CDSS across follow-up visits.

| Patient | Visit | Patient 1 | Patient 2 | Patient 3 |
| --- | --- | --- | --- | --- |
| PANSS<br>(reduction rate) | Pre-surgery baseline | 101 | 121 | 92 |
|  | Post-surgery baseline | 88 (18.3%) | 115 (6.6%) | 70 (35.5%) |
|  | 4-week | 80 (29.6%) | 105 (17.6%) | 68 (38.7%) |
|  | 8-week | 66 (49.3%) | 109 (13.2%) | 65 (43.5%) |
|  | 12-week | 54 (66.2%) | 107 (15.4%) | 62 (48.4%) |
|  | 24-week | 52 (69.0%) | 104 (18.7%) | 49 (69.4%) |
| CGI-S | Pre-surgery baseline | 5 | 6 | 5 |
|  | Post-surgery baseline | 5 | 5 | 5 |
|  | 4-week | 5 | 5 | 5 |
|  | 8-week | 4 | 5 | 5 |
|  | 12-week | 4 | 5 | 5 |
|  | 24-week | 3 | 5 | 4 |
| GAF | Pre-surgery baseline | 35 | 25 | 31 |
|  | Post-surgery baseline | 42 | 25 | 43 |
|  | 4-week | 44 | 27 | 45 |
|  | 8-week | 50 | 21 | 48 |
|  | 12-week | 58 | 30 | 52 |
|  | 24-week | 58 | 25 | 62 |
| CDSS | All visits | 0 | 0 | 0 |

Note: PANSS reduction rate was defined as (pre-surgery baseline score-current score)/(pre-surgery baseline score-30). PANSS, Positive and Negative Syndrome Scale; CGI-S, Clinical Global Impressions-Severity scale; GAF, Global Assessment of Functioning; CDSS, Calgary Depression Scale for Schizophrenia

In an exploratory analysis of longitudinal change, the Friedman test indicated a significant difference in PANSS total score across the eight follow-up assessments (χ² (7) = 19.99, p = 0.0056), accompanied by a substantial effect size (Kendall’s W = 0.952). A permutation-based monotonic trend analysis corroborated a strong decreasing trajectory over time, with a mean within-subject Spearman correlation of -0.964 and a one-sided permutation p < 0.001. These findings are detailed in Table S1 and should be interpreted as exploratory given the limited sample size.

### Neurocognition

At baseline, Patient 1 exhibited the most pronounced global cognitive impairment, with markedly reduced MATRICS Consensus Cognitive Battery (MCCB) neurocognitive and overall composite T-scores. Following DBS, the neurocognitive composite T-score increased from −6 to 18 in Patient 1 and from 39 to 53 in Patient 3 over 24 weeks, while remaining unchanged in Patient 2 (8 at all assessments). Likewise, the overall composite T-score increased from −8 to 13 in Patient 1 and from 36 to 54 in Patient 3 but remained stable in Patient 2 (6-4) (Table S2).

At the cognitive domain level, working memory showed progressive improvement across all three patients. Visual inspection also suggested improvements in speed of processing and reasoning and problem solving in Patients 1 and 3. Improvements in verbal fluency and attention/vigilance were also observed in Patient 1, whereas Patient 3 showed gains in social cognition. In contrast, verbal learning, visual learning, and social cognition showed variable trajectories across individuals.

### DBS programming and optimization

During initial programming, single-contact stimulation produced contact- and amplitude-dependent stimulation-induced responses in all three patients (Fig. 3). The most commonly observed adverse effects were sensory discomfort, transient mood elevation, and increased motor activity. These effects generally occurred at higher stimulation amplitudes and resolved through parameter adjustments or cessation of stimulation. Heart rate showed modest fluctuations during stimulation without clinically significant autonomic adverse events. These acute response profiles informed subsequent DBS programming.

**Fig. 3.**
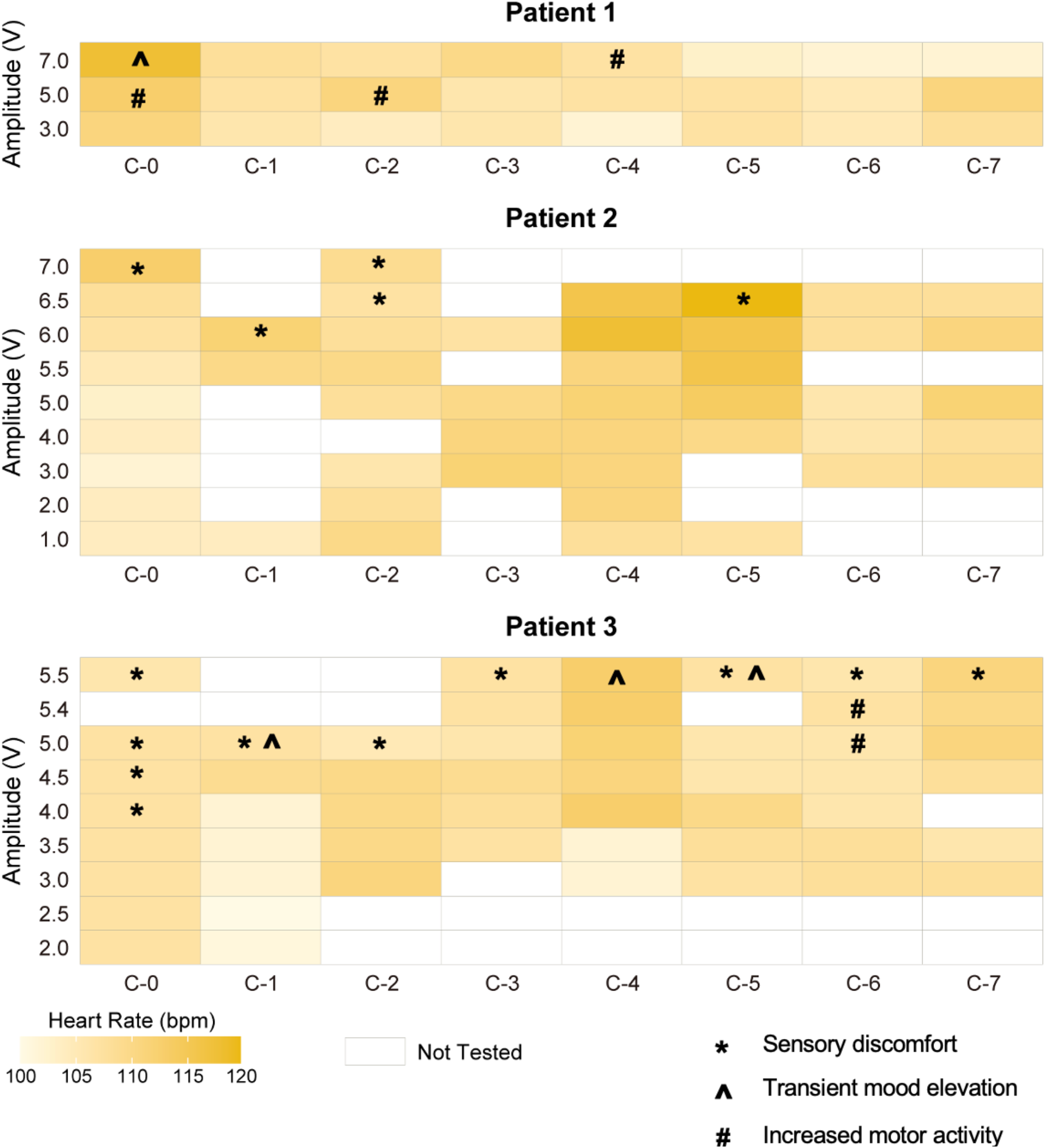
Heart rate responses and stimulation-induced effects during single-contact stimulation testing. Heatmaps depict heart rate (bpm) across stimulation amplitudes and individual contacts for each patient. Symbols denote stimulation-induced sensory discomfort (*), transient mood elevation (^), and increased psychomotor activity (#). Patient 1 demonstrated an elevated mood, spontaneous smiling, and heightened motor activity. Patient 2 reported a sensation of tingling. Patient 3 reported a vibrating sensation in the chest, tingling at the base of the nose, elevated mood, and pressured speech.

DBS parameters were progressively optimized throughout follow-up according to clinical response and tolerability (Table S3). Although initial programming employed conventional parameter settings informed by previous NAc DBS studies, subsequent optimization required patient-specific adjustments in active contacts, stimulation amplitude, pulse width, and daily stimulation schedules. Active contact configuration and stimulation amplitudes were the parameters most frequently modified, whereas pulse width and stimulation frequency remained largely stable.

The extent of optimization varied considerably between patients, with Patient 1 requiring the most complex programming strategy, including differential daytime and nighttime stimulation settings, while Patients 2 and 3 achieved stable therapeutic parameters after fewer adjustments. These observations highlight substantial interindividual variability in both stimulation sensitivity and programming requirements.

### Safety and tolerability of NAc-ALIC DBS

DBS was generally well tolerated throughout the 24-week follow-up (Table S4). No serious adverse events were observed during the follow-up period. There were no surgery-related complications, such as intracranial hemorrhage or infection, and no hardware-related complications, including lead displacement, lead fracture, or implantable pulse generator (IPG) malfunction. No stimulation-related adverse events resulted in treatment discontinuation.

Transient increases in Simpson–Angus Scale (SAS), Abnormal Involuntary Movement Scale (AIMS), and Barnes Akathisia Rating Scale (BARS) scores were noted during the initial postoperative programming period but resolved following stimulation optimization. No persistent or progressive extrapyramidal adverse effects were observed during follow-up.

Notably, after the first week of stimulation, Patient 1 developed mild involuntary movements of the upper and lower limbs (AIMS score = 12; corresponding item severity rated as 2, mild) while receiving stimulation through contacts C0/C2/C4/C6 (210 μs, 145 Hz, 4.5 V; 07:30–22:00, OFF overnight). Stimulation was subsequently reprogrammed by discontinuing NAc contacts and stimulating the ALIC only (contacts C2/C3/C6/C7; 210 μs, 145 Hz, 6.0 V during the daytime and 4.0 V overnight). The involuntary movements resolved completely before the Week 4 assessment, accompanied by normalization of the AIMS score. No other stimulation-related adverse events were reported.

Patient 2 presented with mild antipsychotic-associated extrapyramidal symptoms at baseline (SAS total score 8), including reduced arm swing and gait, mild rigidity, and fine finger tremor. These symptoms gradually improved following adjustment of antiparkinsonian medication, including an increased dose of trihexyphenidyl, and finally resolved during follow-up. Patient 3 experienced no motor adverse effects throughout the study, with SAS, AIMS, and BARS scores remaining at zero across all assessments.

### Intracranial electrophysiological signals

Longitudinal LFPs analysis demonstrated broadly consistent electrophysiological patterns across bilateral DBS contacts. Homologous contacts within bilateral NAc-ALIC regions showed largely concordant longitudinal changes across frequency bands, indicating preserved hemispheric symmetry post-DBS implantation. Across all patients, alpha- and beta-band relative power remained relatively stable over the 24-week stimulation period, with only minor temporal fluctuations.

As shown in Fig. 4, clinical responders (Patients 1 and 3) exhibited a convergent electrophysiological pattern characterized by increased gamma-band relative power and decreased theta-band activity following ALIC-DBS. In contrast, the non-responsive Patient 2 showed a less consistent longitudinal trajectory, lacking the coordinated gamma enhancement and theta suppression observed in responders. These observations raise the possibility that longitudinal changes in low-frequency oscillatory activity, particularly theta-band activity, may be associated with clinical response to ALIC DBS.

**Fig. 4.**
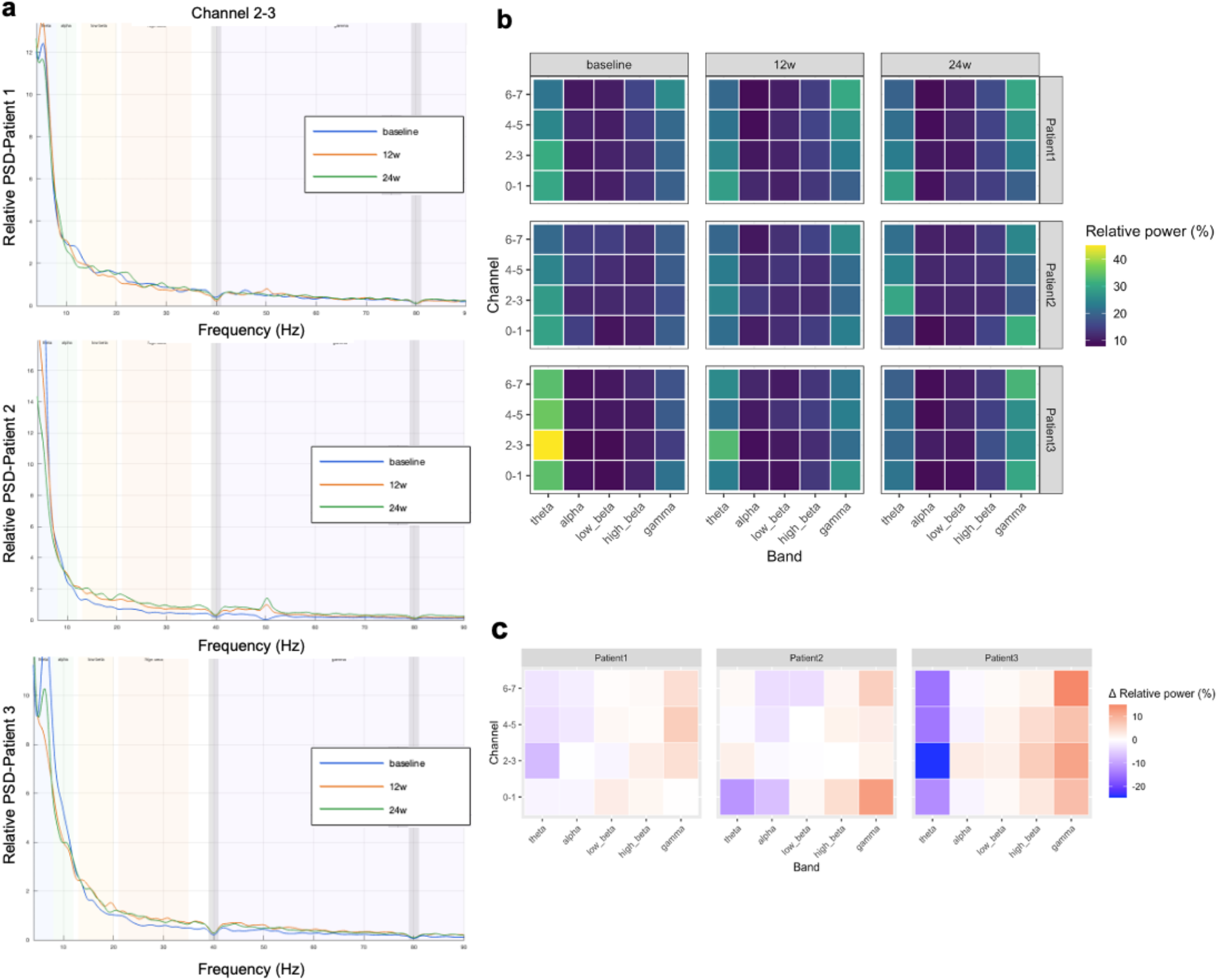
Longitudinal intracranial electrophysiological changes following NAc-ALIC DBS. (a) Relative power spectral density of bipolar channel 2–3 recordings from Patients 1–3 across baseline (blue line), 12-week (red line), and 24-week (green line). (b) Channel × frequency-band heatmap illustrating relative power distribution across recording contacts. (c) Channel × frequency-band heatmap showing longitudinal changes in relative power (Δ relative power) after DBS stimulation.

### Extended observations

Following completion of the prespecified 24-week follow-up, all three participants elected to continue receiving clinical care and were observed beyond the study period. Subsequent outpatient visits and assessments were conducted at the discretion of the participants and their caregivers, without a predefined follow-up schedule.

After 27 weeks of continuous stimulation, the non-rechargeable IPG in Patient 1 approached battery depletion, as indicated by the device warning system. Two weeks after complete battery depletion, the patient’s PANSS total score was 58. By 36 weeks after battery depletion, the PANSS total score had risen to 90, approaching the postoperative baseline level. After evaluating treatment options, Patient 1 and caregivers opted against battery replacement due to financial considerations and subsequently chose to undergo surgical removal of the DBS system.

In Patients 2 and 3, the implanted devices remained in situ after stimulation was discontinued. For Patient 2, stimulation parameters were maintained as set during the 12-week programming session. Following study completion, the clozapine dosage was gradually reduced by a total of 50 mg/day in two steps. After 32 weeks of DBS, the PANSS total score further decreased to 95, corresponding to a 28.6% reduction from the pre-surgery baseline.

For Patient 3, stimulation parameters remained unchanged after the Week 1 programming session. During the extended follow-up period, persecutory and referential delusions became less firmly held, hostility decreased, and the patient was able to obtain part-time employment. These observations were suggestive of continued functional improvement, although the absence of a predefined assessment schedule precludes formal attribution of these changes to ongoing DBS effects.

## Discussion

In this prospective exploratory study, bilateral NAc-ALIC DBS was associated with substantial clinical improvement in two of three patients with highly TRS, whereas the third patient showed more limited improvement. Clinical improvement was accompanied by gains in global functioning and selected cognitive domains, while no serious stimulation-related adverse events occurred. The marked interindividual variability in clinical response and programming requirements, together with convergent changes in intracranial electrophysiology among responders, provides a basis for further investigation of the NAc-ALIC circuit as a therapeutic and physiological target in TRS.

DBS for schizophrenia remains at an early stage compared with its use in movement disorders^30^, TRD^7,8,31,32^ and OCD^5,33,34^, and the optimal target remains uncertain. A variety of target brain regions are under investigation in TRS, including substantia nigra^35^, and hippocampus^36^. The most directly relevant clinical evidence comes from the pilot RCT by Corripio et al., in which NAc and subgenual anterior cingulate cortex DBS were investigated in patients with TRS^9^. Our study extends this work by targeting the NAc together with the adjacent ALIC.

This anatomical strategy is supported by the organization of fronto-subcortical and fronto-limbic circuits traversing the ALIC, as well as by evidence that the ALIC white-matter integrity is altered in schizophrenia. Diffusion imaging studies have reported reduced fractional anisotropy within the bilateral ALIC in schizophrenia, with ALIC white-matter integrity associated with memory performance^28,29^. The NAc may contribute to this therapeutic effect through its role within mesolimbic, motivational and salience-related circuitry^37^. Dopaminergic signaling in the ventral striatum, including the NAc, has long been implicated in the assignment of motivational and attentional salience, and preclinical work has linked NAc dopamine signaling to behavioral processes relevant to psychosis. Accordingly, we interpret the stimulation target as a circuit-level NAc–ALIC region rather than attributing the observed clinical effects to a single anatomical structure.

During DBS programming, acute responses to single-contact stimulation could be broadly classified into sensory discomfort, transient mood elevation, and increased psychomotor activity, resembling a hypomanic clinical presentation. These observations are broadly consistent with a previous case report describing acute frequency-dependent hypomania during DBS programming in a patient with Parkinson’s disease ^38^. In that case, incremental increases in stimulation voltage were accompanied by abrupt changes in mood and behavior, including increased vocalization, interpersonal intrusiveness, rapid speech, elevated affect, and grandiose thought content, which diminished after stimulation was reduced. An alternative explanation is that stimulation of the NAc–ALIC region may recruit neighboring structures involved in motor regulation. The NAc is anatomically adjacent to the caudate nucleus, and current spread from stimulation contacts may therefore engage nearby striatal territories^39^. These observations underscore the importance of contact-specific testing and careful monitoring during programming.

An additional observation was the relatively apparent reduction in PANSS scores already evident at the postoperative baseline in Patient 1 and 3, before prolonged exposure to therapeutic stimulation. One possible explanation is a microlesion effect associated with electrode implantation. Transient clinical improvement following electrode insertion has been documented in DBS surgery, particularly in movement disorders, and has been proposed to reflect local tissue disruption, edema, microhemorrhage, and transient changes in neuronal or axonal activity^40^. Previous DBS studies have reported an association between the magnitude of the early postoperative microlesion effect and subsequent stimulation response^41,42^, although this relationship is not established as a general predictor.

The intracranial electrophysiological findings provide an additional, exploratory dimension. The two clinical responders showed a convergent increase in gamma-band relative power and reduction in theta-band activity, whereas these changes were less consistent in the non-responder. Alpha- and beta-band relative power remained comparatively stable. These findings are potentially relevant because abnormalities in oscillatory synchronization, including gamma-band activity, have been repeatedly implicated in schizophrenia^43^. Preclinical and clinical studies have linked abnormal gamma activity to dysfunction of inhibitory interneuron networks, particularly parvalbumin-expressing GABAergic interneurons^44,45^.

Importantly, our recordings were obtained during stimulation-off periods and at only a small number of longitudinal time points. Thus, the present data cannot determine whether the observed oscillatory changes were directly induced by stimulation, emerged secondarily with clinical improvement, or reflected other time-dependent processes. Future studies using sensing-enabled DBS systems should compare neural activity during stimulation-on and stimulation-off conditions and determine whether candidate oscillatory features change acutely with stimulation, or track longitudinal symptom fluctuations. If reproducible physiological signatures can be identified, they could ultimately support adaptive DBS strategies^46,47^. Closed-loop neuromodulation has already been demonstrated in an individual with TRD, in whom intracranial electrophysiology was used to identify a symptom-linked biomarker and guide stimulation^48^. Our LFP findings should therefore be considered hypothesis-generating.

The cognitive and safety findings provide preliminary information regarding the tolerability of this approach. The two clinical responders showed improvements in overall MCCB performance over 24 weeks, whereas the non-responder remained relatively stable. Because repeated neuropsychological testing, clinical improvement and practice effects may contribute to these changes, the present data cannot establish a direct pro-cognitive effect of DBS. Importantly, no persistent stimulation-related motor adverse effects were observed. The transient involuntary movements in Patient 1 resolved after reprogramming, which is in accordance with previous evidence that non-mood-related and physiological responses were associated with the ventral lead contacts within the NAc-ALIC region^49^.

Several limitations should be considered. First, this was a three-patient, open-label pilot study. The absence of a control group also prevents separation of DBS effects from nonspecific postoperative effects and expectancy-related changes. Second, the implanted system was not 3.0T-MRI-compatible, precluding longitudinal assessment of treatment-related structural or functional network changes using MRI. Third, LFP recordings were obtained at only three time points and during stimulation-off periods, limiting temporal resolution and preventing direct assessment of stimulation-locked electrophysiological effects. Fourth, stimulation parameters were individually modified during follow-up, making it impossible to attribute clinical outcomes to a single stimulation configuration.

## Conclusion

In this prospective pilot study, bilateral NAc–ALIC DBS was associated with substantial symptom improvement in two of three patients with highly treatment-resistant schizophrenia and was generally well tolerated over 24 weeks. The notable heterogeneity in clinical response and programming requirements, along with early postoperative changes and convergent longitudinal alterations in intracranial oscillatory activity among responders, suggests that therapeutic effects may depend on both patient-specific and circuit-specific factors. Future prospective studies incorporating patient-specific connectivity, standardized programming protocols, and longitudinal neural sensing will be important for determining whether NAc-ALIC DBS can provide reproducible clinical benefit in larger population with TRS and whether intracranial physiological features can identify biologically meaningful subgroups of patients.

## Methods

This study was designed as an open-label pilot study. The study protocol was approved by the Ethics Committee of the Shanghai Mental Health Center (No. 2022-91) and Huashan Hospital (No. 2023-929). The trial was registered at an international clinical trial registry (NCT06112067). All study procedures were conducted in accordance with the principles of the Declaration of Helsinki and all participants provided written informed consent prior to enrollment.

### Inclusion and exclusion criteria

Inclusion criteria were: (1) age 18–65 years at the time of written informed consent; (2) DSM-5 diagnosis of schizophrenia confirmed by the Mini-International Neuropsychiatric Interview; (3) illness duration ≥5 years; (4) meeting criteria for treatment-resistant schizophrenia, defined as either: (a) inadequate response or intolerance to ≥2 antipsychotics (excluding clozapine) administered at adequate dose (chlorpromazine-equivalent ≥600 mg/day) and duration (≥12 weeks), or (b) inadequate response or intolerance to clozapine at an adequate dose (≥300 mg/day or plasma level ≥350 ng/mL) and duration (≥12 weeks); (5) at screening and baseline, a Positive and Negative Syndrome Scale (PANSS) total score ≥70, with either ≥1 positive symptom item (P1, P2, P3, P5 or P6) scored ≥4, or ≥3 negative symptom items (N1–N7) scored ≥4, or ≥2 negative items scored ≥5; (6) Clinical Global Impression–Severity (CGI-S) score ≥4; (7) Global Assessment of Functioning (GAF) score ≤60; and (8) provision of written informed consent by patients and their legal guardians after full explanation of the study.

Exclusion criteria were: (1) current clinically significant risk of suicide or violence; (2) ≥20% reduction in PANSS total score between screening and baseline; (3) severe or unstable systemic disease (including cardiovascular, respiratory, hepatic, renal, hematologic, endocrine or neurological disorders); (4) clinically significant abnormalities on physical examination, laboratory testing, electrocardiography or imaging that would preclude participation; (5) prior implantation of devices such as cochlear implants, cardiac pacemakers or defibrillators, prior implantation of similar neuromodulation devices, or surgery within 6 months; (6) contraindications to deep brain stimulation (DBS) or deemed unsuitable for surgery; (7) confirmed HIV infection; (8) pregnancy or breastfeeding, positive pregnancy test at screening, inability to use effective contraception, or planned pregnancy within 3 months; (9) participation in another clinical trial within 3 months before screening; or (10) any other condition deemed unsuitable by the investigator.

### DBS surgery and neuroimaging

For each patient, a safe trajectory targeting the bilateral nucleus accumbens–anterior limb of the internal capsule (NAc-ALIC) was planned using preoperative magnetic resonance imaging (MRI) co-registered with computed tomography (CT). Preoperative surgical planning was performed using Leksell SurgiPlan software (Elekta, Sweden). The patient’s head was fixed using a Leksell G-frame (Elekta, Sweden) under local anesthesia, followed by CT acquisition. The CT images were subsequently co-registered with the preoperative MRI to facilitate stereotactic trajectory planning and target localization.

Because the implanted DBS system was not compatible with 3.0-T MRI, all MRI acquisitions were performed preoperatively. High-resolution structural MRI and diffusion MRI were acquired on a 3.0-T Prisma scanner (Siemens Healthineers, Germany) equipped with a 64-channel head coil. T1-weighted anatomical images were acquired using a three-dimensional sagittal magnetization-prepared rapid gradient-echo (MPRAGE) sequence with the following parameters: repetition time (TR) = 2300 ms, echo time (TE) = 3.5 ms, inversion time (TI) = 1000 ms, flip angle = 9°, field of view (FOV) = 256 × 256 mm², acquisition matrix = 256 × 256, 192 sagittal slices, and an isotropic voxel size of 1 × 1 × 1 mm³.

Diffusion tensor imaging (DTI) was performed using a multiband echo-planar imaging (EPI) sequence with anterior-to-posterior (AP) phase encoding. The acquisition parameters were as follows: TR = 5000 ms, TE = 90.8 ms, flip angle = 90°, FOV = 220 × 220 mm², acquisition matrix = 112 × 112, 74 slices, and an isotropic voxel size of 2 × 2 × 2 mm³. Diffusion-weighted images were acquired with b values of 0 and 1000 s/mm² along 64 diffusion-encoding directions. Additional images with reversed phase-encoding polarity and field maps were acquired to facilitate correction of susceptibility-induced geometric distortions and B0 field inhomogeneity.

All patients underwent bilateral implantation of DBS leads (1242-S, SceneRay) under general anesthesia. Each lead (diameter, 1.27 mm) comprised four 3.0-mm contacts with inter-contact spacings of 2, 4, and 4 mm from ventral to dorsal, designed to simultaneously target the anatomically adjacent NAc and ALIC. Contacts were numbered C0–C3 for the left lead and C4–C7 for the right lead, from ventral to dorsal. The proximal ends of the leads were implanted subcutaneously in the scalp and connected via extension wires (SR1341, SceneRay) to a non-rechargeable implantable pulse generator (IPG; SR1182, SceneRay) placed in the right subclavicular region. Postoperative CT scans were conducted within 24h to ensure successful implantation and to rule out any surgery-related complications such as intracranial hemorrhage.

Preoperative T1-weighted MRI and postoperative CT images were processed using the MATLAB-based Lead-DBS toolbox (version 3.0)^50^ to reconstruct electrode locations. Postoperative CT images were coregistered to preoperative MRI using Advanced Normalization Tools (ANTs) with settings optimized for subcortical structures, and all images were nonlinearly normalized to Montreal Neurological Institute (MNI) space. Brain shifts correction was applied to compensate for intraoperative tissue displacement, followed by manual refinement with WarpDrive to correct residual registration errors. The fused images were used to visualize DBS lead trajectories and target localization.

### Clinical and neurocognitive outcome assessments and DBS programming

All participants underwent a postoperative recovery period of 21±7 days before initiation of DBS stimulation. Baseline assessments were performed on postoperative Day 1, and an initial programming session was conducted after the recovery period to individually optimize stimulation parameters for each patient. To select the appropriate contacts, we utilized subject-specific tractography derived from diffusion tensor imaging (DTI) results, identifying the electrode contacts most closely associated with cortical regions pertinent to the treatment. Parameters included the therapeutic contact configuration, frequency (0-160 Hz), pulse width (60-210 μs), amplitude (0-7 V), and the daily stimulation duty cycle (on/off schedule). In the current study, due to the absence of immediate feedback for psychiatric and cognitive symptoms, as opposed to movement disorders, the initial determination of DBS parameters is predominantly empirical. Researchers adjust these parameters based on acute responses observed during single-contact stimulation testing in DBS programming sessions^51^.

DBS was then delivered continuously over the 24-week treatment period, with contact selection and stimulation voltage adjusted as needed based on individual clinical response. Patients were required to maintain their pre-existing pharmacological treatments throughout the study, with both medication type and dosage kept as stable as possible unless clinical intervention was deemed necessary.

The primary outcome was the change in Positive and Negative Syndrome Scale (PANSS) total score from baseline to 12-week follow-ups after DBS initiation. Secondary outcomes included response rates (≥40% reduction in PANSS total score from baseline), as well as changes in PANSS positive and negative subscale scores, Clinical Global Impression–Severity (CGI-S), Global Assessment of Functioning (GAF), and Calgary Depression Scale for Schizophrenia (CDSS) scores at the same follow-up points.

Neurocognition using the MATRICS Consensus Cognitive Battery (MCCB) was assessed at preoperative baseline and 12-week, 24-week follow-ups after stimulation. Safety and side effect assessment includes patient self-reported discomfort and scales including SAS (Simpson–Angus Scale), AIMS (Abnormal Involuntary Movement Scale) and BARS (Barnes Akathisia Rating Scale). Following each follow-up assessment, DBS programming was conducted to refine stimulation parameters and optimize both clinical efficacy and patient tolerability.

### DBS local field potential acquisition and preprocessing

Local field potentials (LFPs) were recorded from NAc-ALIC deep brain stimulation (DBS) electrodes at baseline, 12-week, and 24-week using the SceneRay LFP recording system. Bipolar derivations were constructed from four adjacent contact pairs (0–1, 2–3, 4–5, and 6–7). Signals were sampled at 1000 Hz under resting-state conditions for approximately 90 seconds per session. To ensure consistency, recordings were obtained immediately after stimulation was temporarily turned off, and continuous time-series data were stored for offline analysis.

All preprocessing and spectral analyses were performed in MATLAB R2023b (MathWorks) using EEGLAB v2022.1 and custom scripts. Detailed procedures are provided in eMethods (Supplement 1). Power spectral density (PSD) was estimated using Welch’s method. Absolute and relative band power were computed within canonical frequency bands: theta (4–8 Hz), alpha (8–12 Hz), low beta (13–20 Hz), high beta (21–35 Hz), and gamma (40–90 Hz). Relative band power was defined as the proportion of total power within the 4–90 Hz range.

### Statistical analysis

Descriptive statistics were presented as means with standard deviations or medians with interquartile ranges for continuous variables, and as proportions for categorical variables, depending on data distribution. Given the small sample size, longitudinal changes in PANSS scores were primarily evaluated descriptively using individual trajectories and percentage improvement from baseline. As an exploratory group-level analysis, Friedman test was used to assess differences across repeated follow-up visits. In addition, a permutation-based monotonic trend analysis was performed to test whether PANSS scores showed a decreasing trend over time. A threshold of P-value <0.05 was set to confirm significance, and 95% confidence intervals were reported to express statistical uncertainty.

## Data Availability

All data produced in the present study are available upon reasonable request to the authors.

## Author Contributions

Liu DT, Wang Z and Xu YF conceived and designed the study. Lu C, Zhai ZL, Xue JX, Zhao YQ, Xiang Q, Zhuo KM, Lei HB and Wang P contributed to recruitment and the overall implementation of the study. Zhang X, Zhuo KM, Lei HB, Wang P, Shen LN and Lang LQ performed the surgical procedure and perioperative management. Lu C, Zhai ZL, Xue JX, Zhao YQ, Xiang Q and Chen L programmed the stimulation parameter of DBS. Shen YF and Han HQ contributed to data monitoring and management. Lu C, Zhang X and Zhai ZL conducted the data analysis and drafted the original manuscript. All authors contributed to data interpretation, critically revised the manuscript for intellectual content, and approved the final version for submission.

## Acknowledgments

The authors sincerely thank all participants and their caregivers for their contribution to this study.

## Conflicts of interest

Dr. Lei Chen is an employee of SceneRay Co. Ltd, Suzhou, the manufacturer of the DBS device used in this study. He provided technical input regarding device operation and critically reviewed the manuscript. The company had no role in study design, data collection, data analysis, interpretation of the results, or the decision to submit the manuscript for publication. The other authors declare no competing interests.

## Funding

This work was supported by the Grant from Shanghai Hospital Development Center (SHDC2022CRT01, SHDC12024131), and the National Natural Science Foundation of China (82371504, 82171496). The founders had no role in the study design, analysis, or in the writing of the report or the decision to submit the report for publication.

## Data sharing statement

Individual participant data underlying this study, including relevant clinical and imaging data, are not publicly available due to patient privacy concerns and institutional restrictions. Data may be made available to qualified investigators upon reasonable request and subject to data use agreements with the corresponding institutions contributing each case.

## Supplement 1. eMethods

### Detailed DBS LFP signal processing and analysis procedures

#### 1. LFP data acquisition

Local field potentials (LFPs) were recorded using the SceneRay DBS system at three time points: baseline, 12 weeks, and 24 weeks. Recordings were obtained from chronically implanted hippocampal DBS electrodes.

A bipolar referencing scheme was applied using four adjacent contact pairs (0–1, 2–3, 4–5, and 6–7) to minimize common-mode noise and volume conduction effects. Signals were sampled at 1000 Hz. Each recording session consisted of approximately 90 seconds of continuous data acquired during resting-state conditions.

To ensure comparability across visits, DBS stimulation was temporarily turned off immediately prior to recording, and continuous raw LFP time series were stored for subsequent offline analysis.

#### 2. Signal preprocessing

All preprocessing was conducted in MATLAB R2023b (MathWorks) and EEGLAB v2022.1.

First, the initial and final 5 seconds of each recording were removed to reduce transient artifacts:

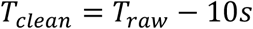

Second, a 1 Hz high-pass Butterworth filter (2nd order, zero-phase) was applied to remove slow drifts and baseline fluctuations. Third, notch filtering was performed to remove narrowband interference at 40 Hz and its harmonics (e.g., 80 Hz), which are commonly introduced by wireless sensing hardware:

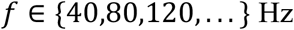

Band-stop filters were implemented using zero-phase filtering to avoid phase distortion.

#### 3. Power spectral density estimation

Spectral power was estimated using Welch’s method. For a signal *x*(*t*), PSD was computed as:

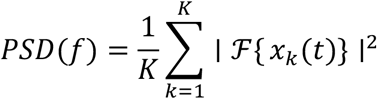

where *x_k_*(*t*)denotes segmented Hanning-windowed epochs.

Window length was set to 1024 samples with zero overlap and FFT length of 1024, resulting in a frequency resolution of:

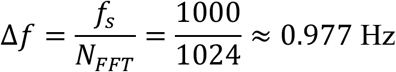

Absolute band power was computed as:

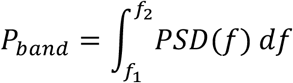

Total power was defined over 4–90 Hz:

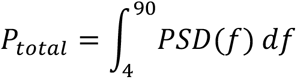

#### 4. Relative band power computation and visualization

To reduce inter-individual variability in impedance and electrode placement, relative band power (RP) was calculated as:

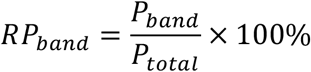

For visualization of spectral trajectories, relative PSD curves were smoothed using a Gaussian kernel (window size = 7 frequency bins).

For each subject and bipolar channel, relative PSD curves were plotted for baseline, 12-week, and 24-week visits to enable longitudinal comparison.

Frequency bands were visualized using semi-transparent shaded regions:

• Theta: 4–8 Hz

• Alpha: 8–12 Hz

• Low beta: 13–20 Hz

• High beta: 21–35 Hz

• Gamma: 40–90 Hz

Notch-filtered frequency ranges (39–41 Hz and 79–81 Hz) were marked using grey shaded areas to indicate potential spectral contamination.

Y-axis scaling was automatically determined using the 99th percentile of pooled spectral values across all visits, with an additional 10% margin to preserve dynamic range while ensuring comparability across time points.

#### 5. Time–frequency analysis

Time–frequency representations were computed using the short-time Fourier transform (STFT):

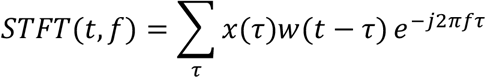

with a 2-second Hanning window and 50% overlap. FFT length was set to 2048 samples. Spectral power was converted to decibels:

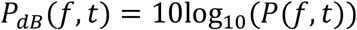

To reduce inter-trial variability, median normalization was applied:

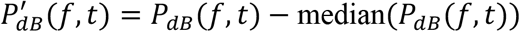

For visualization, STFT maps from baseline, 12-week, and 24-week sessions were plotted side-by-side for each subject and channel.

A common color scale was applied across time points, defined using the 5th and 95th percentiles of the pooled normalized distribution:

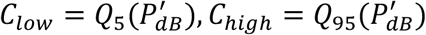

Horizontal reference lines were drawn at 8, 12, 20, 35, and 40 Hz to delineate canonical frequency bands.

Frequency range was restricted to 4–90 Hz, and visualized using the Turbo colormap with individual color bars for each subplot.

## Supplement 2. eResults

**Table S1.**
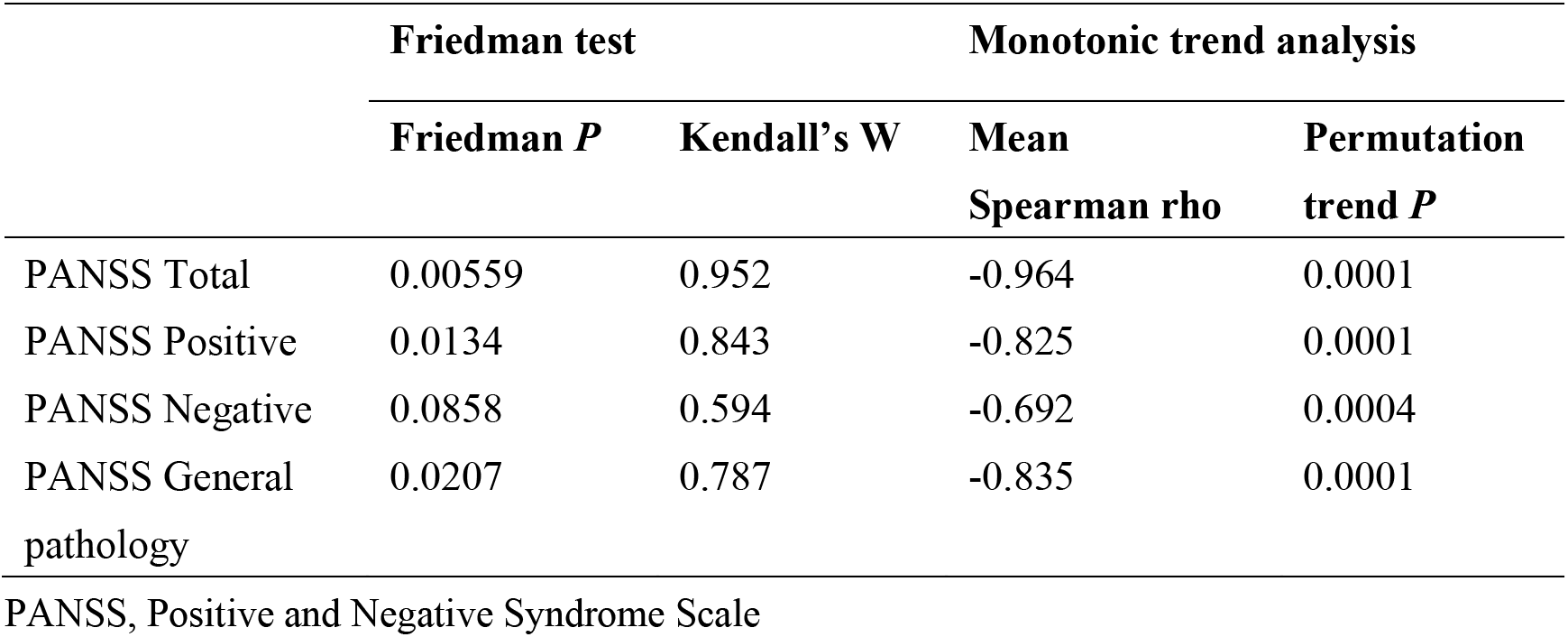
Exploratory group-level analysis of PANSS.

**Table S2.**
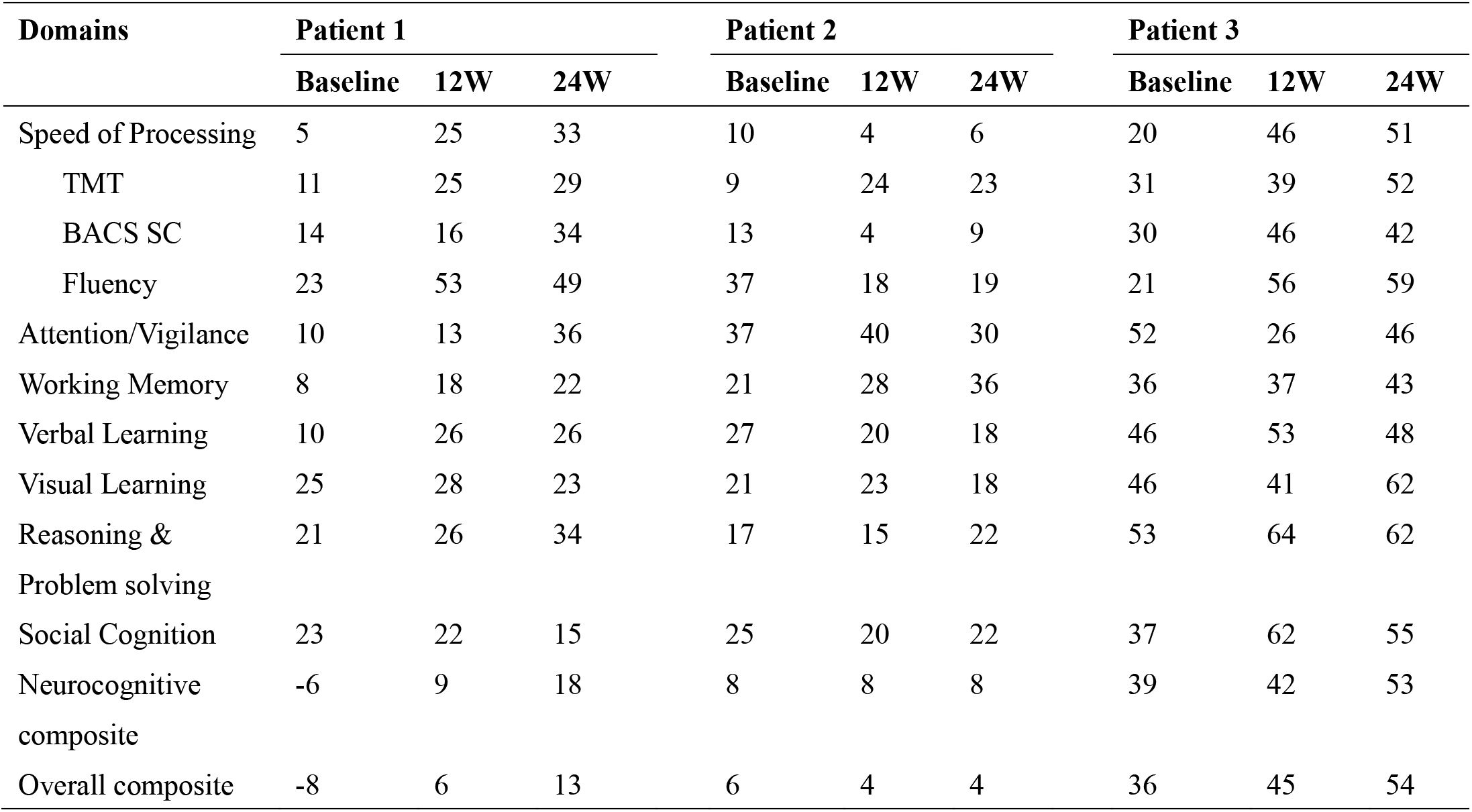
MCCB composite T-scores at pre-surgery baseline, 12-week, and 24-week of DBS treatment.

**Table S3.**
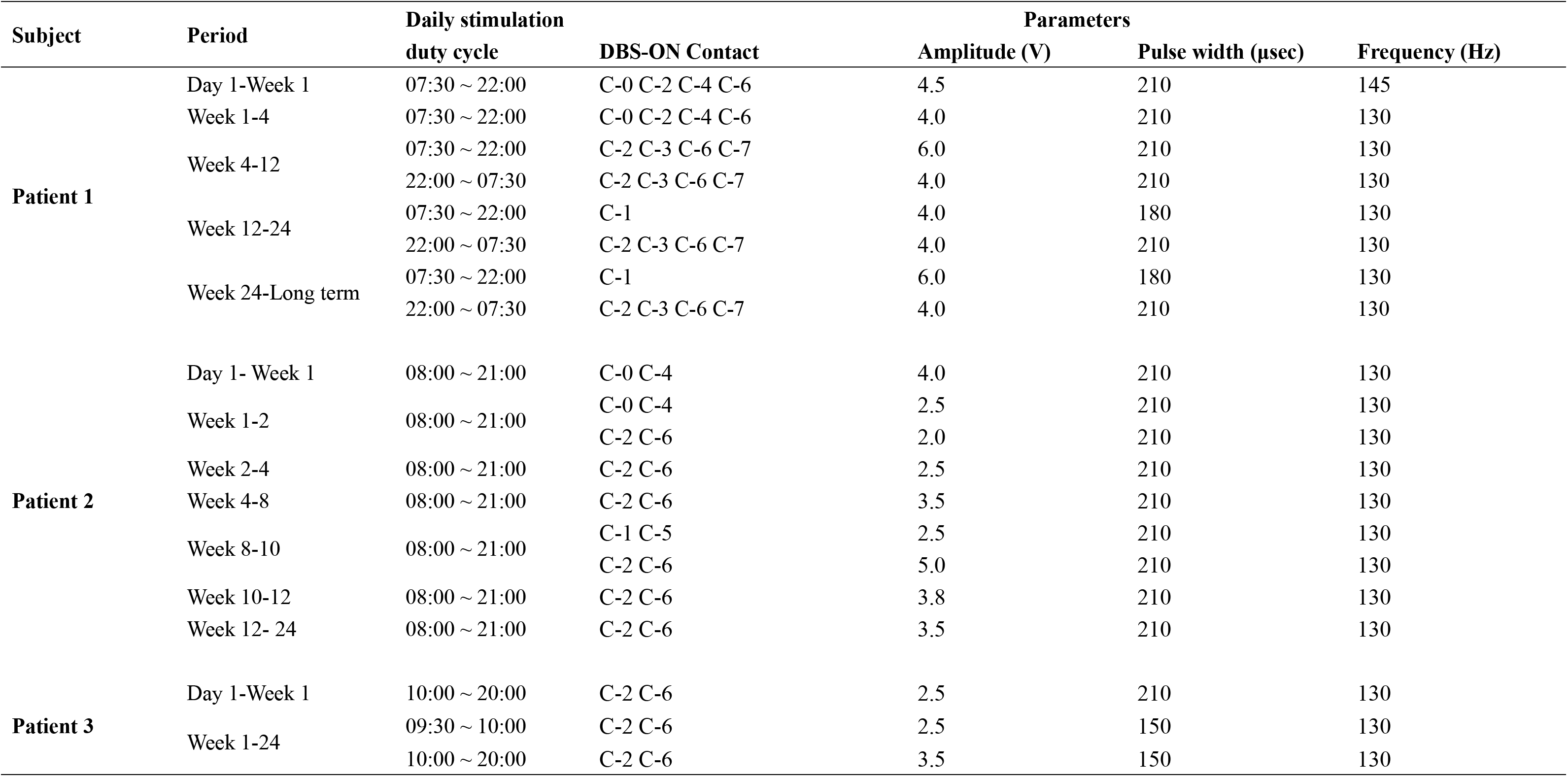
DBS parameter optimization trajectories in Patient 1, 2 and 3.

**Table S4.**
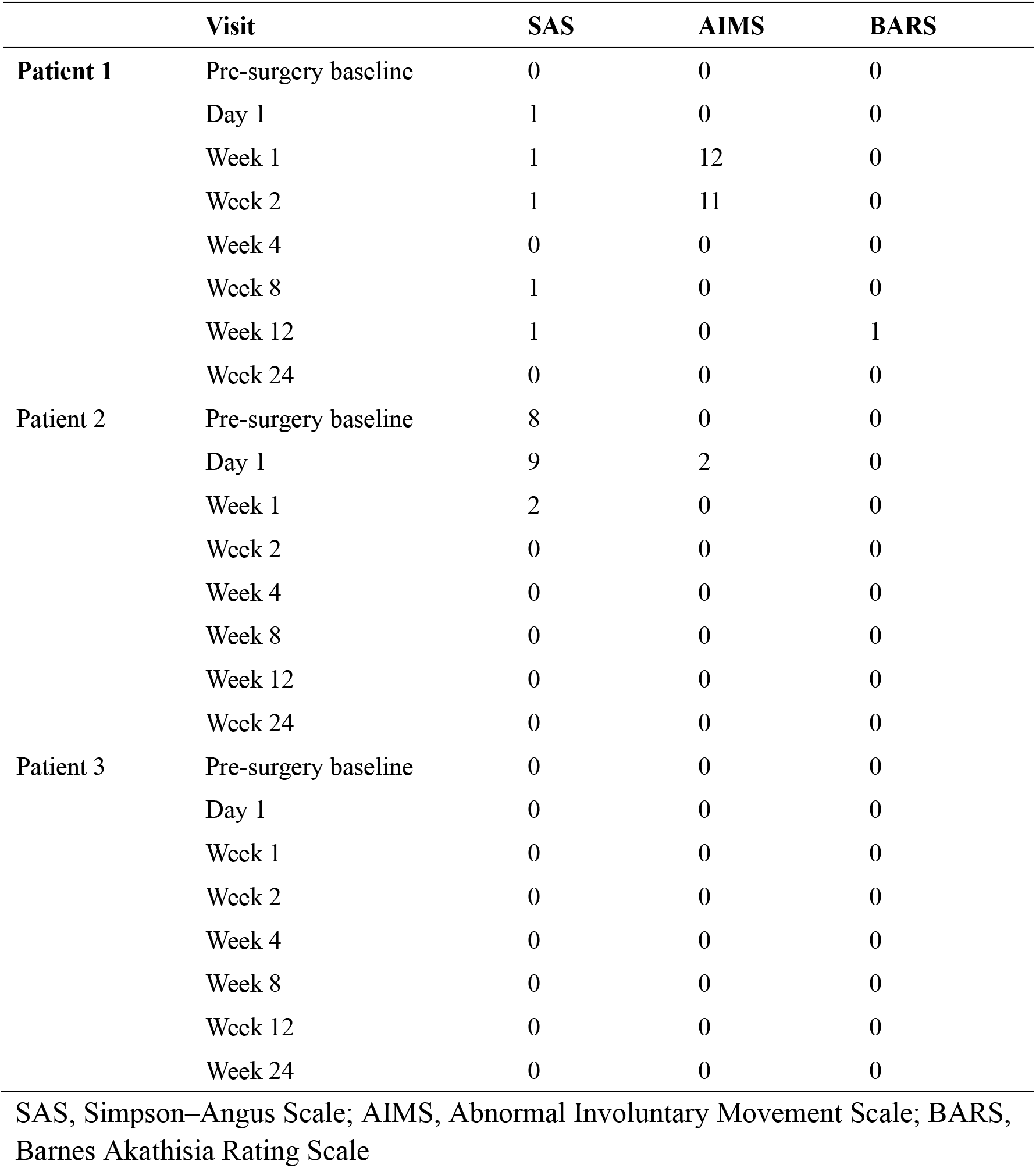
Longitudinal SAS, AIMS, and BARS scores during DBS treatment.

|  | Visit | SAS | AIMS | BARS |
| --- | --- | --- | --- | --- |
| **Patient 1** | Pre-surgery baseline | 0 | 0 | 0 |
|  | Day 1 | 1 | 0 | 0 |
|  | Week 1 | 1 | 12 | 0 |
|  | Week 2 | 1 | 11 | 0 |
|  | Week 4 | 0 | 0 | 0 |
|  | Week 8 | 1 | 0 | 0 |
|  | Week 12 | 1 | 0 | 1 |
|  | Week 24 | 0 | 0 | 0 |
| Patient 2 | Pre-surgery baseline | 8 | 0 | 0 |
|  | Day 1 | 9 | 2 | 0 |
|  | Week 1 | 2 | 0 | 0 |
|  | Week 2 | 0 | 0 | 0 |
|  | Week 4 | 0 | 0 | 0 |
|  | Week 8 | 0 | 0 | 0 |
|  | Week 12 | 0 | 0 | 0 |
|  | Week 24 | 0 | 0 | 0 |
| Patient 3 | Pre-surgery baseline | 0 | 0 | 0 |
|  | Day 1 | 0 | 0 | 0 |
|  | Week 1 | 0 | 0 | 0 |
|  | Week 2 | 0 | 0 | 0 |
|  | Week 4 | 0 | 0 | 0 |
|  | Week 8 | 0 | 0 | 0 |
|  | Week 12 | 0 | 0 | 0 |
|  | Week 24 | 0 | 0 | 0 |

SAS, Simpson–Angus Scale; AIMS, Abnormal Involuntary Movement Scale; BARS, Barnes Akathisia Rating Scale

## Notes

### Clinical Trial

NCT06112067

### Author Declarations

The ethics committee of the Shanghai Mental Health Center, Shanghai Jiao Tong University School of Medicine (No. 2022-91) and the ethics committee of Huashan Hospital, Fudan University (No. 2023-929) gave ethical approval for this work.

